# Design of a computer model for the identification of adolescent swimmers with low BMD

**DOI:** 10.1101/19001792

**Authors:** Jorge Marin-Puyalto, Alba Gomez-Cabello, Alejandro Gomez-Bruton, Angel Matute-Llorente, Alejandro Gonzalez-Aguero, Jose Antonio Casajus, German Vicente-Rodriguez

**Affiliations:** GENUD (Growth, Exercise, NUtrition and Development) Research Group, Universidad de Zaragoza, Zaragoza, Spain; Faculty of Health and Sport Sciences, Huesca, Universidad de Zaragoza, Spain; EXERNET red de investigación en ejercicio físico y salud para poblaciones especiales; Centro Universitario de la Defensa, Zaragoza, Spain; Centro de Investigación Biomédica en Red de Fisiopatología de la Obesidad y Nutrición (CIBERObn), Spain; Instituto Agroalimentario de Aragón (IA2), Universidad de Zaragoza – CITA

**Keywords:** Osteoporosis prevention, decision tree, physical fitness

## Abstract

**Objectives:** This paper aims to elaborate a decision tree for the early detection of adolescent swimmers at risk of presenting low bone mineral density (BMD), based on easily measurable fitness and performance variables.

**Methods:** Bone mineral status of 78 adolescent swimmers was determined using DXA scans at the hip and subtotal body. Participants also underwent physical fitness (upper and lower body strength, running speed and cardiovascular endurance) and performance (swimming history, speed and ranking) assessments. A gradient boosting machine regression tree was built in order to predict BMD of the swimmers and to further develop a simpler individual decision tree, using a subtotal BMD height-adjusted Z-score of −1 as threshold value.

**Results:** The predicted BMD using the gradient boosted model was strongly correlated with the actual BMD values obtained from DXA (r=0.960, p<0.0001) with a root mean squared error of 0.034 g/cm^2^. According to a simple decision tree, that showed a 73.9% of classification accuracy, swimmers with a body mass index (BMI) lower than 17 kg/m^2^ or a handgrip strength inferior to 43kg with the sum of both arms could be at higher risk of having low BMD.

**Conclusion:** Easily measurable fitness variables (BMI and handgrip strength) could be used for the early detection of adolescent swimmers at risk of suffering from low BMD. The presented decision tree could be used in training settings to determine the necessity of further BMD assessments.

**Summary box:** *What are the new findings?:*

- Adolescent swimmers with a low BMI or handgrip strength seem more likely to be at higher risk of having low BMD.
- Subtotal BMD values predicted from our regression model are strongly correlated with DXA measurements.

*How might it impact on clinical practice in the future:*

- Healthcare professionals could easily detect adolescent swimmers in need of a DXA scan.
- The computer-based regression tree could be included in low BMD management and screening strategies.

## INTRODUCTION

Osteoporosis is a metabolic disease that is characterized by a deterioration in skeletal tissue including a clinically low bone mineral density (BMD) and a compromise for the microarchitecture of the bone [1]. Osteoporosis affects 22.1% of women and 6.6% of men over 50 years in the European Union and the total number of patients with this condition is expected to increase in the following years due to demographic variations [2]. This structural fragility entails a lower tolerance to stress which may play a role in up to a 90% of bone fractures [3]. These osteoporotic fractures have been linked to a decrease in the quality of life, to the apparition of disabilities or even mortality [4]. Nowadays, osteoporosis treatment and prevention has become one of the primary concerns for healthcare systems in developed countries [5]. In fact, a total of 33 different clinical practice guides on osteoporosis screening and management issued by institutions all around the globe have been identified and evaluated in a recent systematic review [6], concluding that collaboration and consensus is needed in the elaboration of these guidelines.

Adolescence stands as a decisive period for osteoporosis prevention, since around 40% of adult bone mass is created during this stage [7] and will influence the peak BMD reached in early adulthood. Achieving the highest peak BMD possible is key for the prevention of osteoporosis later in life [7], given that fracture risk is expected to be halved with an increase of one standard deviation in peak bone mass [8].

Physical activity participation and calcium intake are among the controllable factors that are known to affect bone health during childhood and adolescence [9]. Consistent and positive effects on bone development have been found by different reviews focused on sport practice [10,11] and exercise interventions [12]. Additionally, these benefits on bone status obtained from physical activity have been observed to persist in later stages in life [13]. However, not all sport modalities have proven to be beneficial to bone, especially non-weight bearing activities such as swimming may not have a positive effect on bone health [14,15].

Osteoporosis has been defined as a “silent” disease, given that no pain or other symptoms are perceived by the subject who is affected by this condition [3]. For this reason, osteoporosis and low BMD can remain undetected for years and sometimes it is only discovered once the osteoporotic fracture has occurred. There are subjects, thus, who are unknowingly affected by this condition, unaware of their higher risk of fracture and while this condition remains unnoticed no preventive measures could be taken.

Early detection of subjects affected by low BMD is therefore of paramount importance. As such, public health recommendations include an osteoporosis screening for the elderly [16] using the current gold standard method for BMD evaluation, which is the dual-energy X-ray absorptiometry (DXA). However, this clinical evaluation is rarely performed in other population segments, which might also be susceptible from suffering of a decreased BMD such as swimmers.

Correctly identifying the target group for BMD assessments is important regarding the cost-effectiveness of osteoporosis management [17]. However, performing a DXA scan to all adolescent swimmers would not be cost-effective, especially taking into account that the younger the target group, the higher the number of scans needed to prevent one fracture [18]. For this reason, the assessment of different variables related to BMD, more accessible to researchers or healthcare workers, may provide a tool for determining which subjects might need a deeper evaluation of their bone mineral status.

Therefore, the main goal of the present study is to elaborate a screening method for detecting potential risk of low BMD in adolescent swimmers based on easily measurable variables.

## MATERIAL AND METHODS

### Participants and study design

Eighty-six adolescent swimmers (41 females, all Caucasian and aged 10-18 years) participated in the present cross-sectional study, which is part of the broader RENACIMIENTO project [19]. Participants had to have a minimum of 3 years of regional swimming competition experience and train at least 6 hours per week in order to be part of the study. Exclusion criteria included smoking, taking medication known to alter bone and suffering from chronic diseases or musculoskeletal disorders.

Written informed consent from parents was obtained and all participants expressed their agreement. The protocol study was approved by the Ethics Committee of Clinical Research from the Government of Aragón (ref. CP08/2012, CEICA, Spain) and the ethical guidelines for human research outlined by the Declaration of Helsinki (revision of Seoul 2008) were followed.

### Patient and Public Involvement

Participants were not involved in the design, conduct or reporting of the research.

### Anthropometric and bone measurements

Participants underwent the anthropometric examination wearing no shoes and minimal clothing. Height was measured with a stadiometer to the nearest 0.1 cm (SECA 225, SECA, Hamburg, Germany) and weight to the nearest 0.1 kg with an electronic scale (SECA 861, SECA, Hamburg, Germany). Body mass index (BMI) was calculated as weight (kg) divided by squared height (m^2^).

Bone mineral content (g) and areal BMD (g/cm^2^) was determined by means of a dual-energy X-ray (DXA) scan at the whole body and the hip, evaluated with the pediatric version of the QDR-Explorer software, version 12.4 (Hologic Corp., Bedford, MA, USA). All the scans were performed by the same qualified operator who had been trained in the operation of the scanner, the positioning of subjects, and the analysis of scans, according to the manufacturer’s guidelines. Coefficients of variation for the DXA measurements in our laboratory have already been published [20] and were 2.3% for BMC and 1.3% for BMD.

Subtotal (whole body less head) BMD height-adjusted Z-scores were calculated according to the reference values provided by Zemel et al. [21]. A Z-score of −1 was used as the threshold value for the purpose of categorizing subjects with a low BMD. Subtotal whole body was used as it is one of the regions recommended by the International Society for Clinical Densitometry in pediatric populations[22]. Height-adjustment is also advised in this official stand and other publications [22,23].

### Evaluation of pubertal stage

Pubertal maturation was determined by self-assessment of secondary sexual characteristics, with the assistance of a graphical scale, following the method established by Tanner [24], which has been demonstrated as a valid and reliable method to assess sexual maturity among adolescent athletes [25].

### Fitness assessment

Four different components of physical fitness were assessed using field tests. Strength of the upper limbs was determined by the sum of both arms in a maximum isometric handgrip strength with a dynamometer (TKK 5101, Takei Corp., Tokio, Japan), while strength of the lower limbs was assessed by the standing long jump test. Running speed was calculated from the time to complete a 30-m sprint, and aerobic endurance was assessed by means of a 20-m shuttle run test [26]. All tests were performed twice and the best result from both attempts was recorded with the exception of the aerobic endurance test, which was performed once.

### Performance and questionnaires

Swimming history was acquired from a self-reported questionnaire in which participants stated their weekly hours of training and their swimming competition experience (in years). The structured questionnaire also included information of current and past participation in other sports. Additionally, swimming performance was obtained consulting the official timing of swimming competitions, recording the participants’ time in 50-m free-style and their FINA ranking points, an official metric used by the International Swimming Federation to track the performance of swimmers. Daily calcium intake (in milligrams) was calculated from a food frequency questionnaire that included daily, weekly and monthly consumption frequency of various calcium-containing aliments such as cheese or bread [27], which has been validated for adolescent swimmers [28].

### Statistical analysis

Statistical analyses were performed using SPSS for Windows version 22.0 (SPSS Inc., Chicago, IL, USA) with the significance level set at p<0.05. Additionally, the construction of the decision tree was performed with the statistical programming language R [29], including the packages *rpart* [30] and *gbm* [31].

Kolmogorov-Smirnov tests were used to confirm the normality assumption and outlier exploration was performed for all variables included in the study. T-tests for independent samples were used to check the differences between subjects with and without low BMD values on the fitness and performance variables.

### Decision tree modelling

70% of the sample was randomly selected to build a decision tree in order to identify the fitness and performance variables that perform a better discrimination between BMD groups, whereas the remaining 30% of the sample was used to test its classification accuracy. Cross tabulation of sex and pubertal development status across groups was performed and chi-square statistics were used to determine the homogeneity of the group distribution in categorical variables between the training and testing subsamples.

Two decision trees were constructed, following different approaches. The first one was a regression tree (treating hip BMD as a continuous variable) fitting all measured variables. Gradient boosting [32,33] was implemented to increase its precision. From this initial computer model, a second model was developed. This decision tree [34] included solely the nine variables that were significant in the previous model and considered only two possible outcomes; being above or below a Z-score of −1 on the total hip BMD. A summary of the variables included in the decision tree modelling is provided in **table 1**. The code used for the construction of both models can be consulted in **Supplementary Material 1**.

**Table 1.**
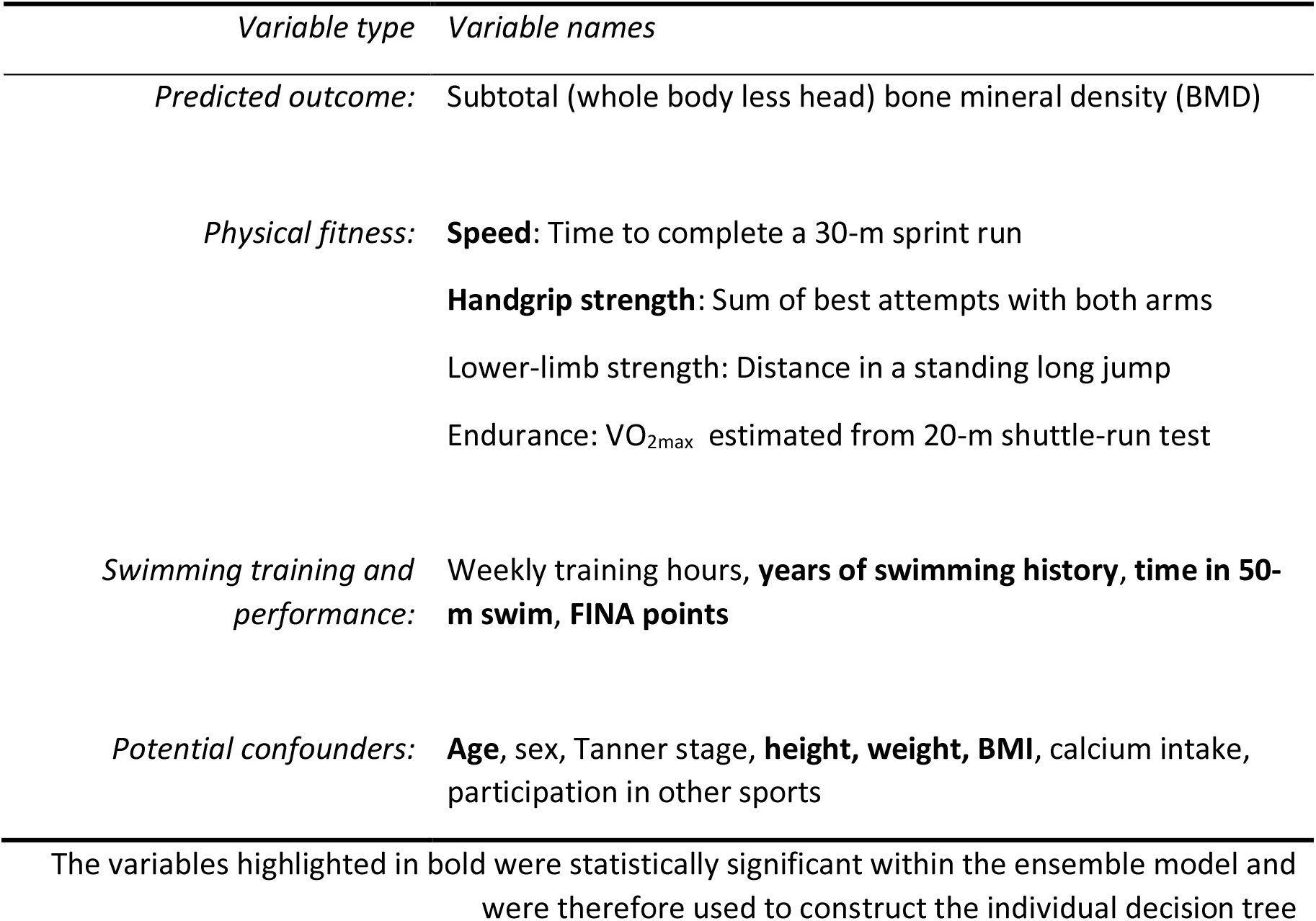
Summary of the variables included in the regression and decision trees

| *Variable type* | *Variable names* |
| --- | --- |
| *Predicted outcome:* | Subtotal (whole body less head) bone mineral density (BMD) |
| *Physical fitness:* | **Speed:** Time to complete a 30-m sprint run<br>**Handgrip strength:** Sum of best attempts with both arms<br>Lower-limb strength: Distance in a standing long jump<br>Endurance: VO<sub>2max</sub> estimated from 20-m shuttle-run test |
| *Swimming training and performance:* | Weekly training hours, **years of swimming history, time in 50-m swim, FINA points** |
| *Potential confounders:* | **Age**, sex, Tanner stage, **height, weight, BMI**, calcium intake, participation in other sports |

The variables highlighted in bold were statistically significant within the ensemble model and were therefore used to construct the individual decision tree

## RESULTS

### Participant characteristics

After removing participants with incomplete or outlier data, 78 participants out of the total sample of 86 swimmers were analyzed. **Table 2** presents their descriptive characteristics, stratified according to their random allocation to the training or testing subsample. No differences between groups were found for any studied variable.

**Table 2.** Descriptive characteristics of the participants

| ***General characteristics and anthropometric variables*** | Overall (n=78) | Training (n=55) | Testing (n=23) |
| --- | --- | --- | --- |
| Sex (male/female) | 40 / 38 | 26 / 29 | 14 / 9 |
| Tanner stage (I/II/III/IV/V) | 2 / 18 / 16 / 38 / 7 | 1 / 10 / 13 / 27 / 4 | 1 / 6 / 3 / 11 / 2 |
| Age (years) | 14.3 ± 1.9 | 14.3 ± 1.8 | 14.3 ± 2.2 |
| Height (cm) | 163.8 ± 12.0 | 163.1 ± 11.3 | 165.4 ± 13.5 |
| Weight (kg) | 54.3 ± 12.1 | 53.3 ± 11.2 | 56.7 ± 13.9 |
| BMI (kg/m<sup>2</sup>) | 20.0 ± 2.5 | 19.8 ± 2.3 | 20.4 ± 2.8 |

| **Bone mineral variables** | Overall (n=81) | Training (n=57) | Testing (n=24) |
| --- | --- | --- | --- |
| Subtotal BMC (g) | 1388 ± 416 | 1350 ± 392 | 1481 ± 465 |
| Subtotal BMD (g/cm<sup>2</sup>) | 0.867 ± 0.113 | 0.858 ± 0.108 | 0.889 ± 0.123 |
| Total hip BMC (g) | 29.9 ± 9.1 | 28.9 ± 8.2 | 32.1 ± 10.6 |
| Total hip BMD (g/cm<sup>2</sup>) | 0.888 ± 0.133 | 0.879 ± 0.136 | 0.907 ± 0.126 |

Categorical variables expressed as frequencies Continuous variables expressed as mean ± standard deviation No significant differences between groups were found

The results from the fitness and performance comparison between participants above and below the threshold in total hip BMD Z-score can be observed in **table 3**. Differences were found between groups for the handgrip strength, long jump, 50-m swim and FINA points (all p<0.05).

**Table 3.** Fitness and performance comparison between subjects with and without low BMD

| **Fitness variables** | Overall (n=78) | Low BMD (n=24) | Not low BMD (n=54) |
| --- | --- | --- | --- |
| Handgrip strength (kg) | 53.5 ± 17.3 | 46.3 ± 16.1 | 56.7 ± 17.0* |
| Long jump (cm) | 185.6 ± 30.4 | 174.3 ± 31.8 | 191.0 ± 28.5* |
| 30-m run (s) | 5.21 ± 0.48 | 5.33 ± 0.42 | 5.15 ± 0.50 |
| VO<sub>2max</sub> (mL/kg/min) | 50.4 ± 5.3 | 49.3 ± 5.0 | 51.0 ± 5.3 |
| **Training and performance variables** | Overall (n=78) | Low BMD (n=24) | Not low BMD (n=54) |
| Weekly training (h) | 10.0 ± 2.1 | 9.8 ± 2.1 | 10.0 ± 2.1 |
| Swimming history (years) | 7.9 ± 2.9 | 7.6 ± 2.7 | 8.0 ± 3.0 |
| 50-m swim (s) | 31.2 ± 3.5 | 32.8 ± 4.4 | 30.5 ± 2.8* |
| FINA points | 354 ± 84 | 317 ± 82 | 370 ± 80* |

Values expressed as mean ± standard deviation \*Significant differences compared to the low BMD group

### Gradient boosting machine regression tree

After scouting and tuning hyper-parameters for the gradient boosting machine (**Supplementary Material 1**), the optimal robustness was found using a total of 5 iterations of the gradient boosting model. Nine of the variables included in the model had a significant influence. An overview of the significant variables is provided in **figure 1**, where it can be observed that height and weight are the variables that contribute the most to the model prediction (41.9 and 18.5% respectively), followed by the handgrip strength (8.4%).

**Figure 1.**
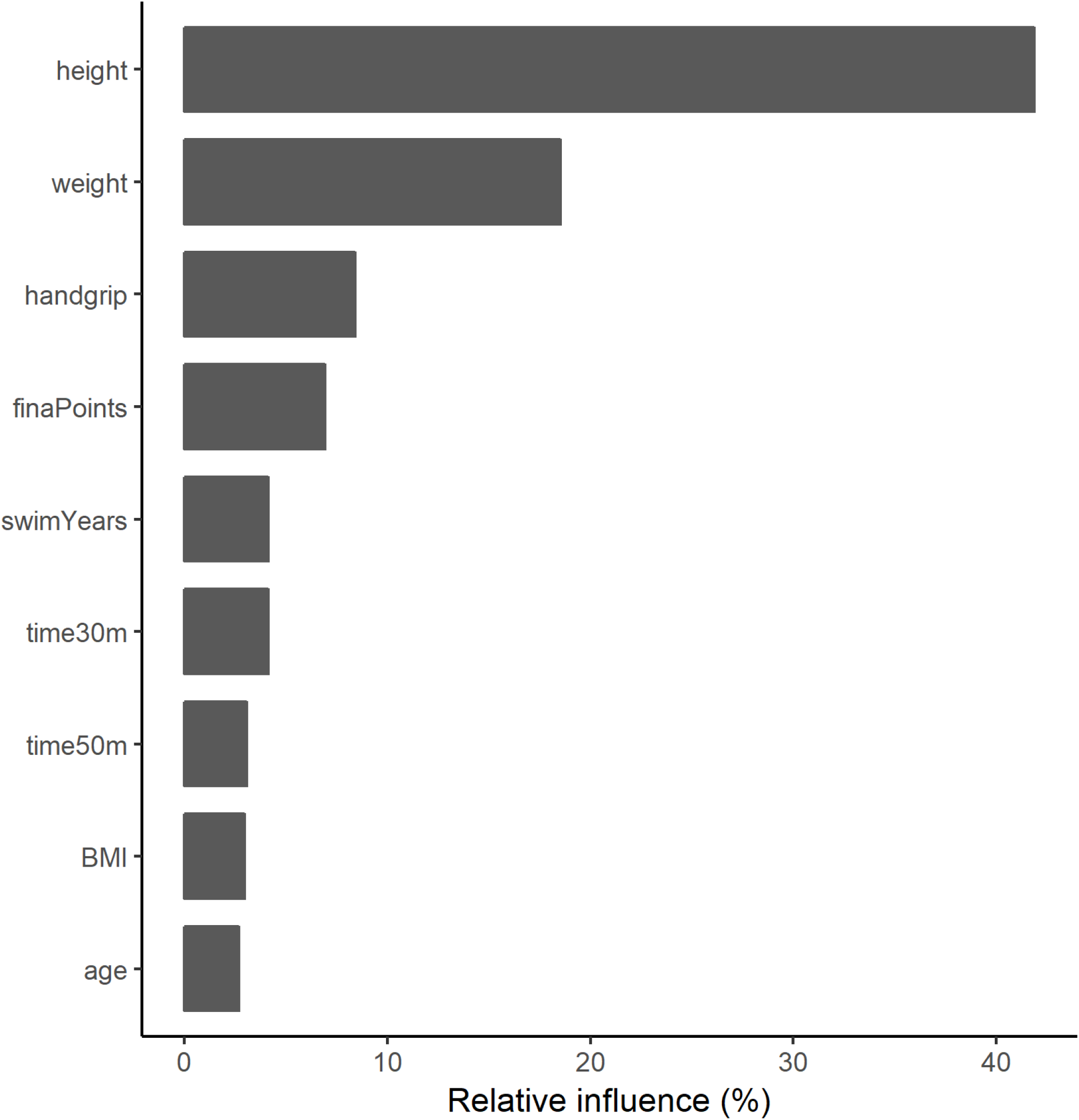
Relative contribution of the main variables of the ensemble model.

When this computer regression model was applied to the 30% of the sample intended to test its prediction accuracy, a root mean squared error of 0.034 g/cm^2^ was obtained. The BMD values obtained from the model were significantly correlated with the actual BMD values as measured by DXA (r=0.960, p<0.0001, **figure 2**). When converted into the correspondent height-adjusted Z-scores, an 73.9% of classification accuracy was reached when comparing participants above and below the proposed threshold.

**Figure 2.**
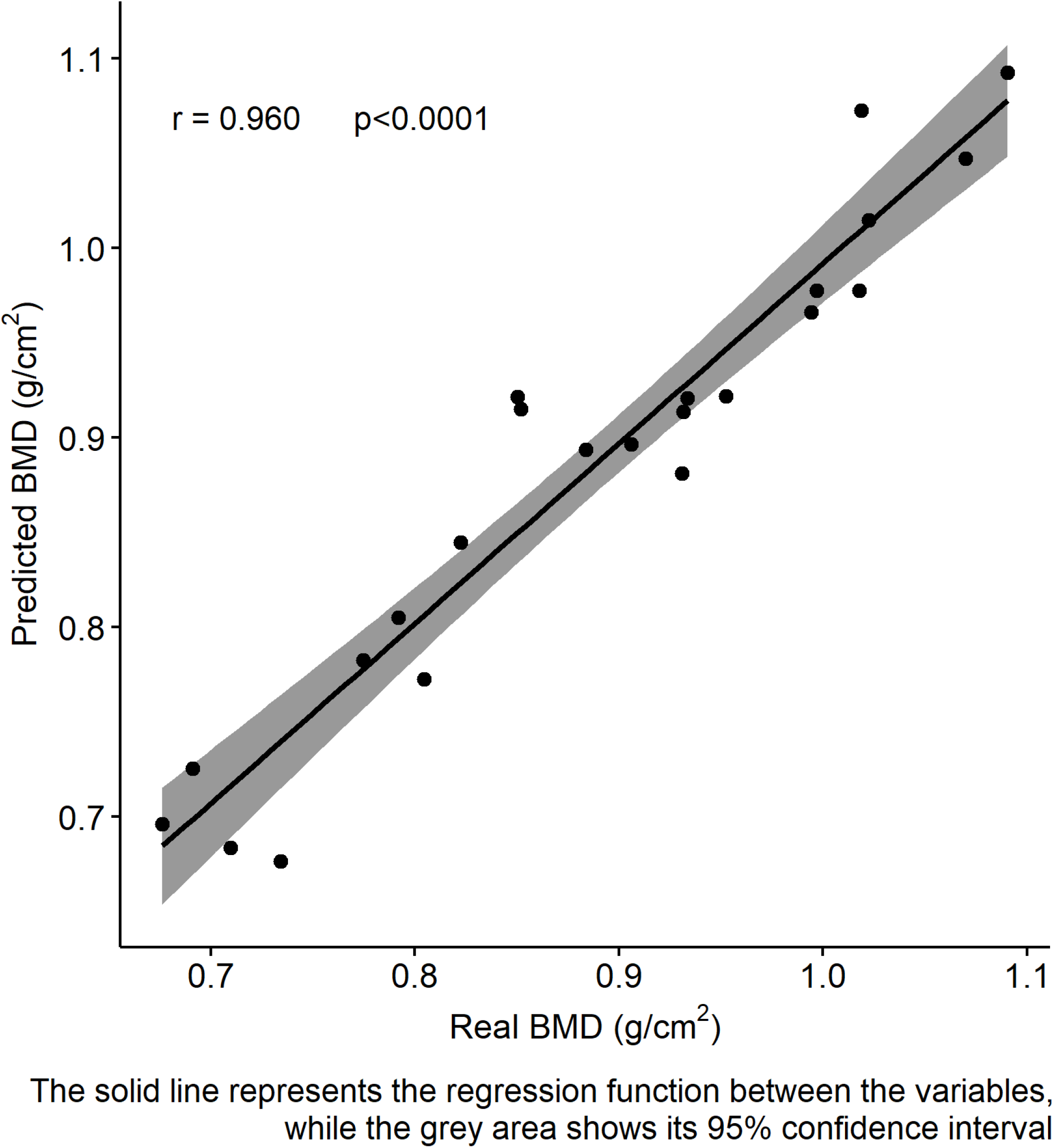
Comparison between the predicted and actual subtotal BMD.

### Individual decision tree

**Figure 3** shows the development of a single classification tree that splits the training sample into four terminal nodes. According to this model, subjects who have either a BMI under 17 kg/m^2^ or less than 43 kg of handgrip strength (summing both arms) present the higher risk of having low subtotal BMD for their age and height. This individual decision tree has an overall classification accuracy of 73.9%, a sensitivity of 50% and a specificity of 82.4%.

**Figure 3.**
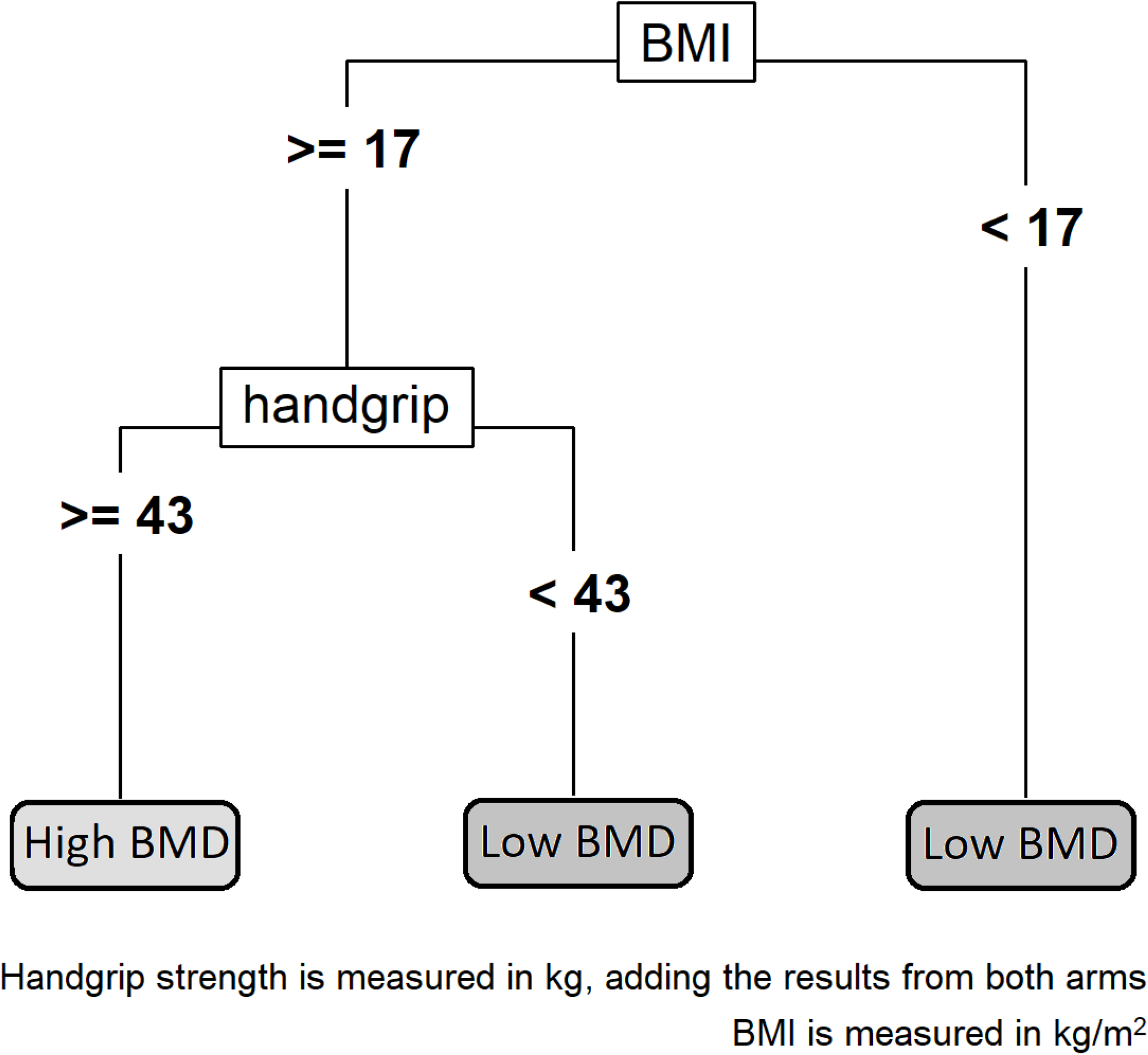
Individual decision tree.

## DISCUSSION

### Main results and relevance

The main results from the present document are that physical fitness and performance variables can be used for the prediction of low BMD in adolescent swimmers and establish the appropriateness of further body composition examinations.

The relevance of the present study is the development of a practical tool for an initial screening of adolescent swimmers at risk of suffering from low BMD. The swimming trainers or healthcare workers can easily measure all the variables included in the model without demanding requirements of material or human resources. Most variables can be measured with the help of a questionnaire or a chronometer, being the main exception the handgrip test, which requires a dynamometer. However, the use of this test is justified for three reasons, since it has been related to other health and performance parameters [35,36], it contributes significantly to both the regression and decision models and it should also be noted that it is a particularly quick, cheap and simple test.

Regarding the specific results of the gradient boosting machine model, height and weight of the participants were the variables that affected the most to their BMD. However, some physical fitness and performance variables accounted for some of the model prediction, especially handgrip strength. The mean squared error of the model did not result in a systematic or proportional bias in the prediction of the total hip BMD. The classification accuracy of 0.739 is not perfect but it is better than chance alone and similar than the areas under the curve (AUC) reported [37] for other screening strategies devised for postmenopausal women such as the FRAX tool [38] (AUC: 0.60), the Simple Calculated Osteoporosis Risk Score [39] (AUC: 0.72) and the Osteoporosis Self-Assessment Tool [40] (AUC: 0.73).

These results are obtained from the model that includes all sixteen variables in a gradient boosting machine and can provide interesting theoretical information. However, this model might not be easy to implement, since it requires a high number of measurements and a computer evaluation of the model. In order to offer a useful tool for trainers that might not have the time and skills needed to perform the complete evaluation, an individual decision tree is provided, which is based solely on the handgrip test and BMI measurement.

In the particular case of our testing sample, three out of the six participants that actually had a subtotal BMD Z-score below −1 would have been recommended to undergo a DXA scan based on the results from our model. Also, only two of the other seventeen participants of the subsample would have been erroneously advised to have their BMD checked.

The accuracy and validity of both the gradient boosting model and the individual decision tree have been assessed with the perspective of trying to simultaneously optimize both the sensitivity and the specificity of the model. However, decision tree analysis allows penalizing differently false positive and false negative cases, by assigning their specific costs. This might be interesting, since it could be argued that in this particular it might be preferable to have a healthy subject scanned rather than failing to identify a participant at risk of having low BMD. A cost-effective analysis of the osteoporosis prevention system could reveal the actual cost of a false negative over a false positive that would in turn provide the optimal balance between specificity and sensitivity that the model should seek. However, doing so would require specific evaluation of the costs of healthcare interventions, which may vary between countries and would therefore limit the geographical applicability of the model.

It is important to note that the purpose of the presented models is not to provide clinical diagnostic or to replace the need DXA scans in any way, but rather is to complement them, serving as a previous screening filter for detecting those subjects who would benefit the most from a DXA assessment. The cost-effectiveness of the osteoporosis screening protocol for postmenopausal women has been confirmed [17]. However, the final implementation of these protocols is not always comprehensive, and sensitization strategies have been implemented [41,42]. A previous study that evaluated the effects of the implementation of a temporary clinical case finding strategy for osteoporosis detection in postmenopausal women showed a great improvement of the screening policy using a simple index based solely on the age and weight of the subject [43]. Additionally, model-based tools have been successfully implemented to assess fracture risk in the elderly in the framework of clinical osteoporosis management [38]. However, to the best of our knowledge, no similar studies have been performed with adolescent swimmers. We consider that this is of primary importance, given the popularity of this sport nowadays in the young populations, and that it will permit the early identification of individuals with low BMD, and it will allow to start taking action early and act preventively in order to avoid future cases of osteoporosis.

### Limitations and future research

Some limitations have to be acknowledged as well. The reduced size and the wide age range of the test subsample might require the confirmation of the validity and adjustment of the model in specific populations. However, it should be pointed out that no differences in age or sex distribution were found between the testing and training subgroups or among the low and normal BMD groups. Additionally, even though fitness and nutrition variables were considered, there might be other variables that affect BMD included in the regression algorithm. Future research should confirm the applicability of the model in other samples as well as to investigate if the addition of other easily measurable variables could improve the prediction accuracy.

### Conclusion

In conclusion, this study presents a theoretical model and a practical tool for early detection of adolescent swimmers at risk of low BMD that can be quickly applied by swimming trainers or health professionals that work as a first step to seek clinical advice to those subjects that were previously unaware of their condition.

## Data Availability

Data files can be requested to the corresponding author

## Acknowledgements and funding

We would like to thank the participants, their families and the coaches for their collaboration in this project. This work was supported by the Spanish “Ministerio de Economía y Competitividad” “Plan Nacional I+D+i2008–2011 (Project DEP2011-29093)” and by “Ministerio de Educación y Ciencia” (Project DEP2005-00046). This project has been co-financed by “Fondo Europeo de Desarrollo Regional” (MICINN-FEDER). JMP received a Grant FPU014/04302 from ‘Ministerio de Educación, Cultura y Deportes’.

~~~
*#loading necessary packages*
library(dplyr)
library(rpart)
library(rpart.plot)
library(gbm)
library(ggplot2)
library(vip)
library(pdp)
library(boot)
library(ggpubr)
*#loading data and isolating the variables included in model construction*
load(“RENACIMIENTO.RData”)
treeDB<-renacimiento[,c(12,2:9,30:37)]
*#for reproducibility*
set.seed(1)
*#randomly selecting 70% of the sample*
random_index<-sample(1:nrow(treeDB),nrow(treeDB))
treeDB<-treeDB[random_index,]
train<-sample(1:78,round(78*0.7,0))
*#creating a matrix to test different hyper-parameter values*
hyper_grid <-expand.grid(
shrinkage = c(.01, .1, .3),
interaction.depth = c(1, 3, 5),
n.minobsinnode = c(2, 5, 8),
bag.fraction = c(.5, .75, 1),
optimal_trees = 0,
min_RMSE = 0
)
*# grid search*
for(i in 1:nrow(hyper_grid)) {
*# reproducibility*
set.seed(123)
*# training model*
gbm.tune <-gbm(
 formula = totalBMD ∼ .,
 distribution = “gaussian”,
 data = treeDB[train,],
 n.trees = 100,
 interaction.depth = hyper_grid$interaction.depth[i],
 shrinkage = hyper_grid$shrinkage[i],
 n.minobsinnode = hyper_grid$n.minobsinnode[i],
 bag.fraction = hyper_grid$bag.fraction[i],
 train.fraction = .75
)
*# adding min training error and trees to grid*
hyper_grid$optimal_trees[i] <-which.min(gbm.tune$valid.error)
hyper_grid$min_RMSE[i] <-sqrt(min(gbm.tune$valid.error))
}
*#checking best fits*
hyper_grid %>%
dplyr::arrange(min_RMSE) %>%
head(10)
*#fine-tuning hyper-parameter based on previous results*
hyper_grid_fine <-expand.grid(
 shrinkage = c(.1, .2, .3),
 interaction.depth = c(3,5,7),
 n.minobsinnode = c(2,3,4,5),
 bag.fraction = c(.5,.625,.75),
 optimal_trees = 0,   # a place to dump results
 min_RMSE = 0    # a place to dump results
)
*# new grid search*
for(i in 1:nrow(hyper_grid_fine)) {
*# reproducibility*
set.seed(123)
*# training model*
gbm.tune <-gbm(
 formula = totalBMD ∼ .,
 distribution = “gaussian”,
 data = treeDB[train,],
 n.trees = 100,
 interaction.depth = hyper_grid_fine$interaction.depth[i],
 shrinkage = hyper_grid_fine$shrinkage[i],
 n.minobsinnode = hyper_grid_fine$n.minobsinnode[i],
 bag.fraction = hyper_grid_fine$bag.fraction[i],
 train.fraction = 0.75
)
*# adding min training error and trees to grid*
hyper_grid_fine$optimal_trees[i] <-which.min(gbm.tune$valid.error)
hyper_grid_fine$min_RMSE[i] <-sqrt(min(gbm.tune$valid.error))
}
*#checking new best fits*
hyper_grid_fine %>%
 dplyr::arrange(min_RMSE) %>%
 head(10)
*#building single model with the identified parameters*
set.seed(123)
boost.fit<-gbm(totalBMD∼.,
   distribution=“gaussian”,
   data=treeDB[train,],
   n.trees=50,
   interaction.depth = 3,
   n.minobsinnode = 3,
   bag.fraction=0.75,
   shrinkage=0.3,
   cv.folds=5)
*#checking model results*
boost.fit
*#plotting model performance*
png(filename = “cvError.png”,width=1600,height=1600,units=“px”,res=300)
gbm.perf(boost.fit, method = “cv”)
dev.off()
*#plotting relative importance of variables*
png(filename = “relativeImportance.png”,width=1600,height=1600,units=“px”,res=300)
par(mar = c(5, 8, 1, 1))
vip::vip(boost.fit,n.trees=50,num_features=9)+theme_classic()+ylab(“Relative influence (%)”)
dev.off()
*#predicting risk in test data to include them in the database*
pred<-predict(boost.fit,n.trees=50,newdata=treeDB[-train,])
*#loading database with the predicted and actual BMD values and its correspondent Z-score*
load(“predMat.RData”)
*#calculating root mean squared error*
sqrt(mean((predMat$realBMD-pred)^2))
*#showing the accuracy of the prediction*
table(real=!predMat$realAdjZ>-1,pred=!predMat$predAdjZ>-1)
*#building single tree for practical application*
indTreeDB<-renacimiento[,c(38,2:9,30:37)]
indTreeDB<-indTreeDB[random_index,]
indTree<-rpart(risk∼height+weight+finaPoints+handgrip+swimYears+time30m+time50m+BMI+age,
   data=indTreeDB[train,],method=“class”,minsplit=20,
   parms=list(loss=matrix(c(0,1,1,0),byrow=TRUE,nrow=2)))
*#plotting contents of the individual tree*
png(filename = “individualTree.png”,width=1600,height=1600,units=“px”,res=300)
rpart.plot(indTree,box.palette=gray(seq(.6, 1, length.out=9)),type=5,extra=0)
dev.off()
*#calculating the prediction on the remaining 30% of the sampling*
indPred<-predict(indTree,newdata=indTreeDB[-train,])[,2]>1/2
*#showing the accuracy of the prediction*
table(real=!predMat$realAdjZ>-1,pred=indPred)
~~~

